# Therapeutic acute intermittent hypoxia alters supraspinal neural control of lower limb movement in multiple sclerosis

**DOI:** 10.64898/2026.11.01.26366948

**Authors:** Hannah R Johnson, Chendong Zou, Ramyar Taha, Jordan N Northrop, Neha A Reddy, Aravind Nehrujee, Alexander Barry, Rachel Kalvakota, Leah O’Shea, Bruce A Cohen, Alexander J Nemeth, Milap S Sandhu, Molly G Bright

## Abstract

Acute intermittent hypoxia (AIH) therapy is a promising neurorehabilitative intervention with well- documented improvement in motor function in people with spinal cord injury. Its effect on lower-limb motor control in multiple sclerosis (MS), where walking impairment is a leading source of disability, remain uncharacterized, and the supraspinal effects of AIH therapy are unknown. In a sham-controlled, double-blind crossover trial (NCT05164705), 23 adults with MS completed 5 sessions of AIH therapy and of sham therapy; magnetic resonance imaging (MRI) was performed before and after each therapeutic intervention. At each timepoint, blood oxygenation level-dependent (BOLD) functional MRI was collected during a repeated plantar- and dorsi-flexion task. Task-evoked activation was modeled at the group level and compared across timepoint and therapeutic arm. Activation changes differed for the left- and right-foot task. AIH augmented activation in the basal ganglia, thalamus, and cortical motor regions during left-foot movement with similar but sub-threshold patterns for the right foot. Right-foot movement additionally showed enhanced deactivation in prefrontal, parietal, and temporal cortices, although these effects were primarily in white matter voxels, limiting their interpretability. These findings suggest that changes in subcortical activation may contribute to motor function improvements following AIH therapy, implicating the basal ganglia-thalamo-cortical motor loop as a target of AIH-induced neuroplasticity.

These results also indicate that AIH therapy modulates supraspinal motor circuitry, supporting further investigation of the cortical and subcortical effects of this emerging therapeutic treatment.

## 1 Introduction

Acute intermittent hypoxia (AIH) is an emerging rehabilitative therapy that drives neuroplastic changes and supports motor recovery in the damaged central nervous system^1^. It consists of brief, repeated exposures to low oxygen (∼9% O_2_) alternating with periods of normoxia. The hypoxic stimulus activates chemoreflex-driven serotonergic neurons in the raphe nuclei, triggering synthesis of brain-derived neurotrophic factor (BDNF) and enhancing motoneuron excitability^2,3^. AIH therapy has been shown to improve walking speed and endurance, joint strength, and dexterity in persons with spinal cord injury, and there is growing interest in expanding the clinical applications of this promising therapeutic intervention^2,4–10^. Investigations are needed to determine whether similar mechanisms may promote functional recovery in further populations with impaired motor pathways, including persons with multiple sclerosis (PwMS).

Multiple sclerosis significantly impairs motor function as demyelination and neuroaxonal inflammation disrupt communication between the central nervous system and the musculature^11^. Persons with multiple sclerosis experience muscle weakness, pain, spasticity, abnormal reflexes, and poor fine motor control, contributing to difficulty maintaining balance and gait and executing everyday tasks^11,12^. Approximately 75% of PwMS have difficulty walking due to sensorimotor impairments^11,12^. Current treatment strategies for multiple sclerosis focus primarily on suppressing inflammation to mitigate disability progression and strategies to manage symptoms. Rehabilitation approaches, including physical and occupational therapies and adaptive devices, have been used for decades to help maintain mobility and quality of life, yet these treatment strategies do not target the neuroplastic mechanisms underlying lasting motor function recovery^13–15^. Given that AIH appears to modulate neural transmission in corticospinal circuits involved in motor control, this therapy presents a promising and distinct method of promoting motor function recovery in PwMS, complementary to existing treatments^16,17^.

Our group has recently presented preliminary evidence that a single session of AIH therapy improves motor function in PwMS, showing increased ankle strength, walking ability, and cognitive processing^18^. However, relatively little is known about *how* AIH alters the underlying neural control of movement in these individuals^18,19^. While lesions and neurodegeneration may impact the integrity of the corticospinal tract in multiple sclerosis, the disease typically involves more diffuse pathology across the brain and spinal cord^20^. Thus, while the mechanisms underlying AIH efficacy may overlap between SCI and MS cohorts, this therapy may act through distinct supraspinal pathways in PwMS. Functional magnetic resonance imaging (fMRI) provides a noninvasive method of studying neural activity, and when combined with a task, can be used to probe changes in sensorimotor activation^21–23^. Previous fMRI studies have shown that patterns of brain activation during voluntary movement are altered in PwMS, with increases in the magnitude and extent of activation compared to healthy subjects, suggesting compensatory activation and functional reorganization of the brain^24–26^. Understanding how AIH therapy alters neural activation associated with motor tasks in PwMS could provide insight into the mechanisms through which this promising treatment improves motor function, facilitating its translation into clinical rehabilitative practice.

We therefore investigated whether repeated sessions of AIH changed neural activation associated with a lower-limb motor task in PwMS. We conducted a randomized sham-controlled crossover trial, performing motor-task fMRI at the beginning and end of five sessions of daily AIH (or sham AIH) therapy. We hypothesized that compensatory neural activation associated with the lower limb motor task would be reduced following AIH therapy.

## 2 Methods

This study was approved by the Institutional Review Board at Northwestern University (STU00219535) and written, informed consent was provided by all subjects before any study procedures took place. Study activities were performed at the Shirley Ryan AbilityLab and Northwestern University. Twenty-seven individuals with relapsing-remitting or secondary-progressive multiple sclerosis were recruited for this study. To be included in the study, participants must have an expanded disability status scale (EDSS) score between 3 and 6.5 and a motor functional system scale (FSS) score between 2 and 4, be between the ages of 18 and 75, free of relapses for at least the past year, safe to be scanned with MRI, and if prescribed dalfampridine, taking the same daily dose for at least two months prior to enrollment.

Individuals with active MS lesions, chronic uncontrolled hypertension or sleep apnea, a history of epilepsy or chronic obstructive pulmonary disease, or who were pregnant were ineligible.

### 2.1 Study Design

A randomized, double-blinded, sham-controlled crossover study design was employed. Participants completed two five-day arms, separated by a washout period of at least four weeks. The order of interventions was randomized. Functional magnetic resonance imaging and motor function assessments were performed on Day 1 and Day 5 of each intervention week, and AIH or sham AIH was delivered on Days 1-5 (Figure 1A). Scanning occurred at a different site to the intervention sessions and motor function assessments, and participants were transported between the two sites in a wheelchair with the support of a clinical aide.

**Figure 1.**
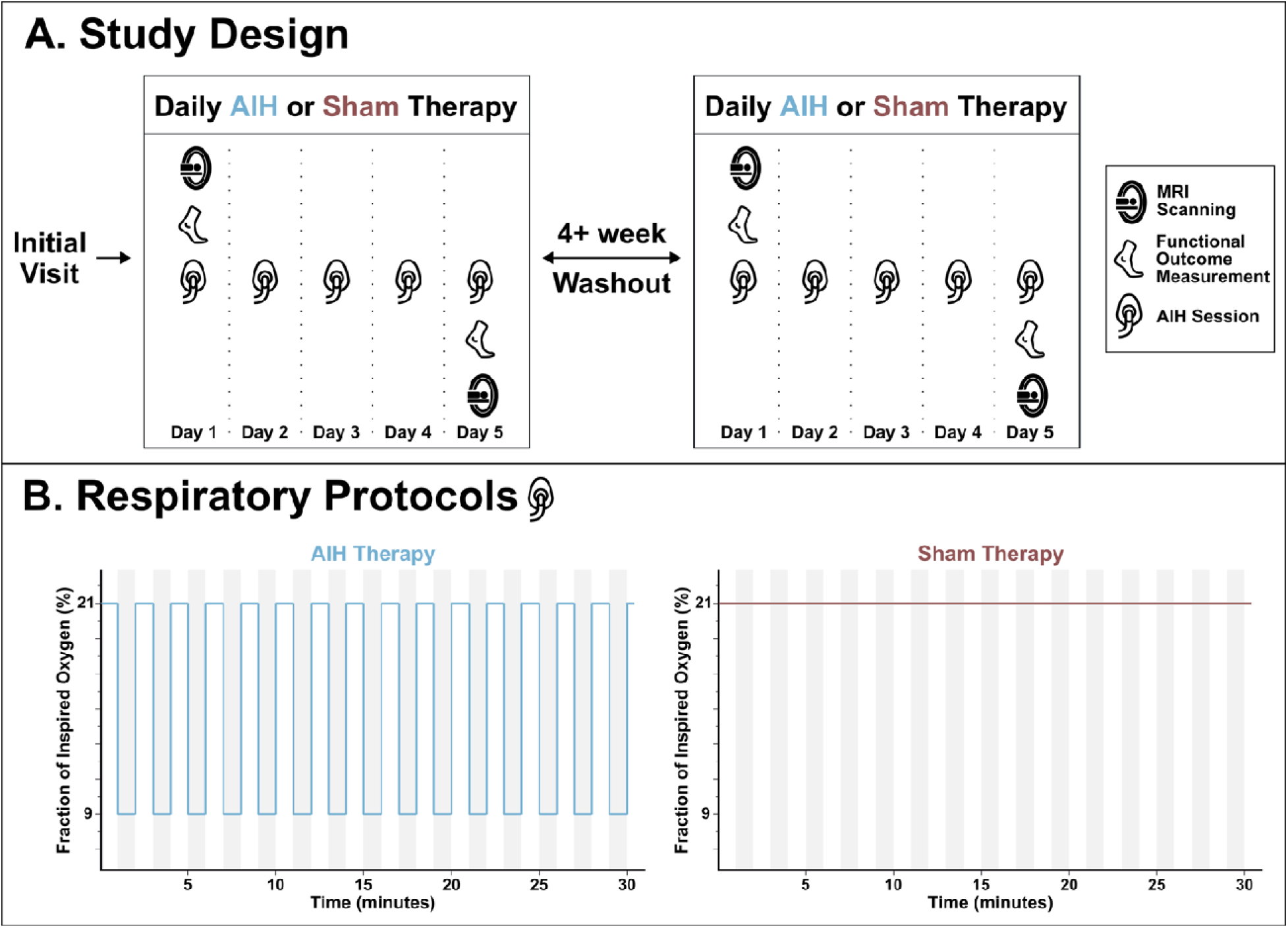
Study Design. Overall study design is shown in Panel A; following an initial screening visit, participants completed two study arms in a randomized order, separated by at least four weeks. Each study arm consisted of five days of daily acute intermittent hypoxia (AIH) or sham therapy. Before AIH or sham therapy on Day 1 and after therapy on Day 5 of each arm, MRI scanning was performed and functional outcome measures were collected. The target respiratory protocols for AIH and sham therapy are shown in Panel B, representing the oxygen content of the air inhaled across time. Grey shading indicates when participants were breathing air from an altitude generator; the generator was calibrated to output hypoxic (9% O_2_) air for AIH therapy sessions and normoxic (21% O_2_) for sham therapy sessions.

### 2.2 Intermittent Hypoxia Protocol

Participants underwent 5 sessions each of AIH and sham AIH, with interventions delivered in the morning on consecutive days within each arm. Participants were seated and fit with a non-rebreathing face mask through which gas mixtures were delivered. The intermittent hypoxia protocol (Figure 1B) lasted approximately 30 minutes and consisted of 15 bouts of hypoxia (9-10% FiO_2_) or sham hypoxia (21% FiO_2_) alternated with bouts of normoxic room air (21% FiO_2_). Hypoxic bout durations were determined individually in each session to achieve a target arterial oxygen saturation (SpO_2_) of 84-88%. Normoxic intervals lasted at least 60 seconds to ensure that participants’ SpO_2_ recovered to at least 92%. Hypoxic and sham hypoxic air were generated by a hypoxia generator (Hyp-123, Hypoxico Inc), and an oxygen monitor continuously monitored the fraction of inspired oxygen of generated gas mixtures.

Oxyhemoglobin saturation and heart rate were continuously monitored (Masimo Rad-57, Danaher; Nonin Model 7500 pulse oximeter, Nonin Medical Inc.) and blood pressure was taken before and after each session (Welch-Allyn Connex, Hillrom).

### 2.3 Imaging Protocol

fMRI was performed on Day 1 and Day 5 of each intervention arm. Scanning was performed on a Siemens 3T Prisma scanner using a 64-channel head coil. A structural multi-echo T1-weighted MPRAGE was acquired with scan parameters^27^: TR = 2170.0 ms, TEs = 1.69/3.55/5.41 ms, FA = 7°, FoV = 256 x 256 mm^2^, voxel size = 1.0x1.0x1.0 mm^3^. fMRI data were acquired with a 2D T2*-weighted multi-band, multi-echo gradient echo echo-planar imaging sequence: TR = 1500.0 ms, TEs = 10.60/27.83/45.06/62.29/79.52 ms, FA = 70°, multi-band factor = 4, GRAPPA acceleration factor = 2, FoV = 210x210 mm^2^, voxel size = 2.5x2.5x2.5mm^3^, phase encoding direction A>>P. A reverse-phase encode sequence (P>>A) was acquired prior to functional scans to aid in distortion correction.

Physiological monitoring was performed throughout scanning at a sampling rate of 100 Hz (PowerLab 8/35 and LabChart, ADInstruments): end-tidal carbon dioxide and oxygen were monitored via nasal cannula and gas analyzer (ADInstruments), breathing via respiration belt (BIOPAC), and pulse via a fingertip pulse transducer (ADInstruments). Participants performed repeated ankle flexion during the fMRI acquisition, with a wedge pillow positioned under their knees for comfort. Visual instructions prompted participants to perform fourteen 16-second blocks of unilateral dorsi- and plantar-flexion, paced at 0.5 Hz, with 10–18 seconds of rest between blocks, and alternating sides for each consecutive block.

Participants were given the chance to practice this task outside of the scanner before each session to ensure comfort and understanding.

### 2.4 Data and Statistical Analysis

#### 2.4.1 Structural MRI Pre-Processing

Subject T1-weighted images were processed with (*fsl_anat,* FSL)^28^, which performs bias-field correction and brain extraction, and registered to the MNI152 standard-space T1-weighted 2mm template image using FSL *flirt* and *fnirt*^29^, which perform linear and non-linear registration, respectively.

#### 2.4.2 Functional MRI Pre-Processing

Data were pre-processed using tools in the FSL and AFNI software suites. The first 10 volumes of fMRI data were removed for each echo to allow for magnetization stabilization. Data were distortion-corrected (*topup*, FSL)^30,31^ and realignment was performed to correct for head motion, registering data from the first echo to a single-band reference image taken at the start of the functional acquisition (*3dvolreg*, AFNI) and applying realignment parameters to subsequent echoes (*3dAllineate*, AFNI). Images were brain-extracted (*bet*, FSL)^32^.

*Tedana* (version 24.0.2)^33,34^ was used to calculate the optimal combination of the multi-echo data and to perform subsequent multi-echo independent component analysis (ME-ICA). The resulting components were automatically classified and accepted or rejected as noise based on the tedana *demo_taskexternalregressors_model* decision tree, as described by Reddy et al.^35^ The optimally combined dataset was smoothed with a 3mm full-width at half maximum Gaussian kernel (*3dmerge,* AFNI) and converted to signal percent change.

#### 2.4.3 Generation of Regressors

Left- and right-foot task regressors were generated by convolving task block timings with the canonical hemodynamic response function. End-tidal peaks in CO_2_ timeseries were automatically detected in MATLAB and manually inspected. Peaks were interpolated to create the end-tidal CO_2_ trace, which was convolved with the canonical hemodynamic response function, rescaled, and demeaned. The demeaned trace was down-sampled to match the temporal resolution of the fMRI timeseries, forming the end-tidal CO_2_ regressor.

#### 2.4.4 Subject-Level Modeling

Functional data for each subject and session were modeled from the signal percent change maps using a general linear model of the left- and right-foot task regressors, the end-tidal CO_2_ regressor, the six motion parameters and their derivatives generated by head-motion realignment, Legendre polynomials up to the fourth order, and rejected ME-ICA components orthogonalized using the conservative method described by Moia et al. and validated for motor task fMRI by Reddy et al.^35,36^

#### 2.4.5 Motor Task Group Analysis

Activation maps from each subject and session were transformed to standard space, applying concatenated transforms from functional to anatomical and anatomical to standard spaces (*flirt* and *fnirt,* FSL). Group-level activation analysis was performed with only data from pre-intervention scan sessions (n=46 from 23 subjects): AFNI’s *3dMEMA* was used to run a one-sample test for the data from each foot separately, with outliers modeled and no covariates included^37^. Resulting group maps were thresholded at p<0.001 and cluster-level α<0.05 (bi-sided, NN=1) after estimating spatial autocorrelation and determining minimum cluster size (*3dFWHMx, 3dClustSim, 3dClusterize,* AFNI).

#### 2.4.6 Group Analysis of the Intervention Effect

AFNI’s *3dANOVA3* was used to determine the group-level impact of the sham and AIH intervention on task activation and significant differences between the two interventions for each foot. A three-way mixed-effects ANOVA was run (*3dANOVA3*), modeling timepoint (pre and post) and intervention (AIH and sham) as fixed effects and subject as a random effect. Minimum cluster size was determined from estimated spatial autocorrelation and generated ANOVA results were thresholded and clustered at α<0.05 (bi-sided, NN=1); p<0.001 was used for activation maps and p<0.05 was used for changes in activation (*3dFWHMx, 3dClustSim, 3dClusterize,* AFNI). Details and locations of significant clusters were summarized with AFNI’s *3dClusterize*, *whereami*, and *3dBrickStat*, using the CA_N27_ML atlas to localize clusters and the Harvard-Oxford subcortical structural atlas’ white matter mask thresholded at 50%.

## 3 Results

Twenty-seven individuals enrolled in the study. Four subjects discontinued study participation due to scheduling conflicts. Twenty-three subjects completed both treatment arms, and their data were included in analyses. Two subjects’ (12 and 20) final scans were not collected due to technical challenges.

Demographic information for all enrolled subjects is shown in Table 1.

**Table 1.** Participant Demographics. Subjects that withdrew prior to treatment are indicated in italics. Treatment order 1 indicates that sham treatment was delivered first; treatment order 2 indicates that AIH was delivered first. M: male; F: female; W: white; AA: African American; >1: more than one race; RR: relapsing-remitting; SP: secondary progressive; R: right; L: left.

| **Study ID** | **Age Range (yrs)** | **Sex** | **Race** | **MS Type** | **EDSS** | **Weaker Side** | **Time Since Diagnosis (yrs)** | **Treatment Order** |
|---|---|---|---|---|---|---|---|---|
| **01** | 50-59 | F | W | RR | 6.5 | R | 13 | 2 |
| **02** | 60-69 | F | AA | RR | 6.5 | L | 30 | 1 |
| **03** | 40-49 | F | W | RR | 3 | L | 9 | 1 |
| **04** | 70-79 | M | AA | SP | 6.5 | L | 2 | 2 |
| **05** | 70-79 | M | W | SP | 5.5 | L | 6 | 1 |
| ***06*** | *40-49* | *F* | *AA* | *RR* | — | — | *8* | — |
| **07** | 50-59 | M | W | SP | 6.5 | L | 24 | 2 |
| **08** | 30-39 | F | W | RR | 6 | L | 6 | 2 |
| **09** | 50-59 | F | AA | RR | 5.5 | L | 14 | 1 |
| **10** | 50-59 | F | >1 | RR | 5 | R | 16 | 1 |
| ***11*** | *50-59* | *M* | *W* | *RR* | *5.5* | *L* | *26* | — |
| **12** | 70-79 | M | W | RR | 3.5 | L | 36 | 2 |
| **13** | 70-79 | M | >1 | RR | 5 | R | 45 | 1 |
| **14** | 30-39 | M | W | RR | 5.5 | R | 3 | 2 |
| ***15*** | *70-79* | *F* | *W* | *RR* | *4.5* | *L* | *21* | *2* |
| ***16*** | *60-69* | *F* | *W* | *SP* | *6.5* | *R* | *16* | — |
| **17** | 50-59 | F | W | SP | 6 | R | 36 | 1 |
| **18** | 50-59 | F | AA | RR | 6.5 | L | 22 | 1 |
| **19** | 60-69 | M | W | RR | 6 | L | 32 | 2 |
| **20** | 50-59 | F | W | RR | 3 | L | 44 | 1 |
| **21** | 70-79 | F | W | SP | 6 | R | 27 | 2 |
| **22** | 50-59 | M | W | SP | 6 | L | 18 | 1 |
| **23** | 60-69 | F | W | SP | 6 | R | 20 | 2 |
| **24** | 60-69 | F | >1 | SP | 6 | L | 32 | 2 |
| **25** | 40-49 | F | AA | RR | 6 | R | 8 | 2 |
| **26** | 60-69 | M | W | RR | 3 | L | 11 | 1 |
| **27** | 60-69 | M | W | RR | 4.5 | R | 41 | 1 |

### 3.1 Neural Activation with Task

Clusters of BOLD signal changes associated with the lower limb task, calculated from pre-intervention sessions, are shown in Figure 2. For both the right and left-foot tasks, significant positive activation was detected in the contralateral primary and supplemental motor cortices, contralateral primary somatosensory cortex, and bilateral opercular, insular, and secondary somatosensory cortices. In the basal ganglia, clusters of significant BOLD signal increases were found in contralateral caudate, contralateral pallidum, and bilateral putamen and superior lateral thalamus. Significant decreases in BOLD signal during the motor task were identified bilaterally in the precuneus, lateral primary somatosensory cortex, posterior parietal cortex, and medial inferior frontal pole. These regions largely align with expected activation as seen in healthy controls, but insular, opercular, and basal ganglia activation may reflect compensatory activation patterns^38–41^. Additionally, while deactivation is expected due to default mode network (DMN) suppression, decreased BOLD signals in the posterior parietal lobe extend beyond the expected DMN regions^42^.

**Figure 2.**
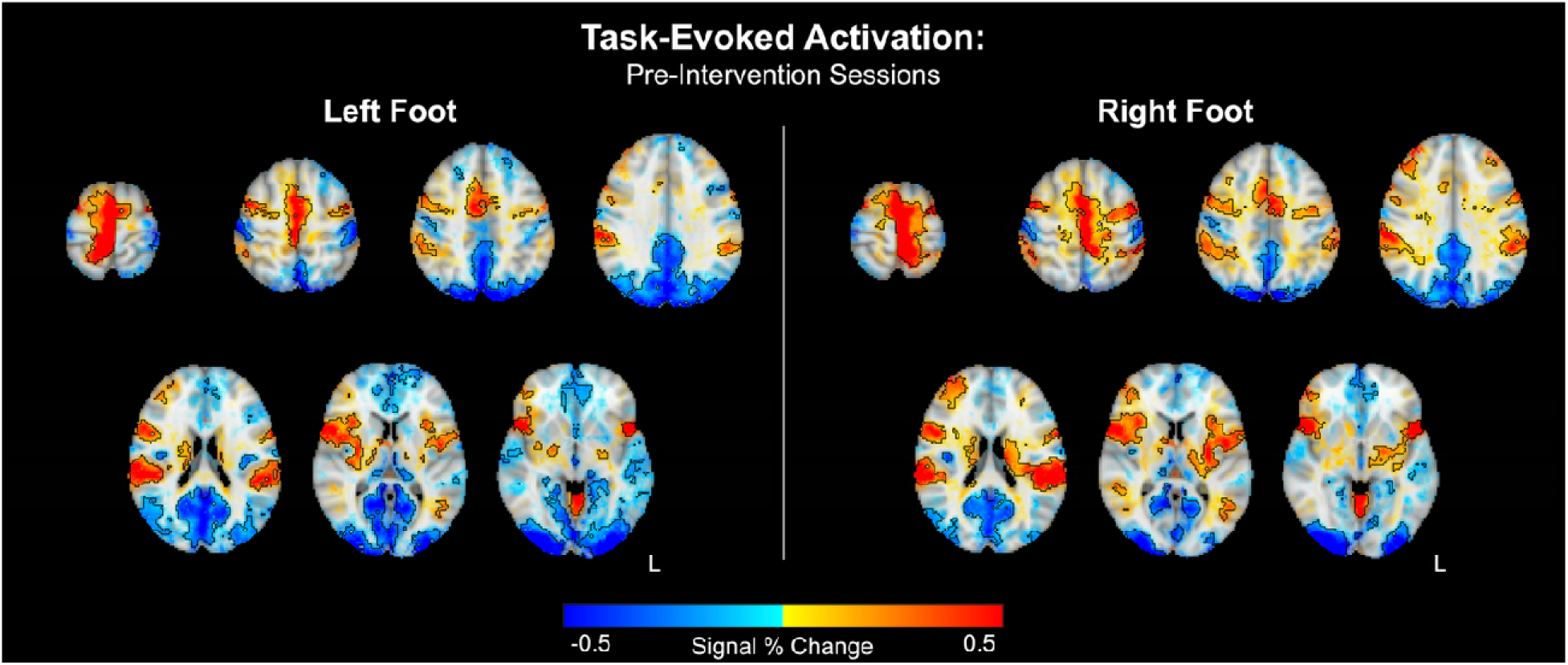
Task Activation. Group maps of task activation were calculated from pre-intervention MRI sessions using BOLD signal changes in response to repeated plantar- and dorsi-flexion with the left and right feet. Signal change opacity is quadratically modulated by the t-statistic^43^. Clusters of significant activation (p<0.001) are outlined in black.

### 3.2 Effect of Intervention on Task Activation

Task activation before and after sham and AIH interventions, as well as the difference in task activation between the timepoints for the left foot are shown in Figure 3. Following the sham intervention arm, negative activation in the precuneus and ipsilateral angular gyrus during the motor task was significantly increased (i.e. these regions became less negative). Significantly decreased activation was seen in the bilateral primary and supplementary motor cortices, insula, and prefrontal cortices, as these regions became negative in the post-intervention scans; however, these clusters are largely comprised of white matter (65-94%) and should therefore be interpreted cautiously. Following the AIH intervention arm, activation significantly increased in the contralateral occipital cortex stemming from a decrease in negative BOLD signal in the post-intervention scan compared to pre-intervention. Sub-threshold decreases in activation were also present in the bilateral prefrontal cortices, similar to the sham condition, and sub-threshold increases were present bilaterally in the thalamus, putamen, pallidum, and caudate resulting from increases in positive BOLD signal. Additionally, changes approaching significance were seen in the bilateral opercular cortex in both sham and AIH interventions: activation decreased with the sham intervention but increased with AIH therapy.

**Figure 3.**
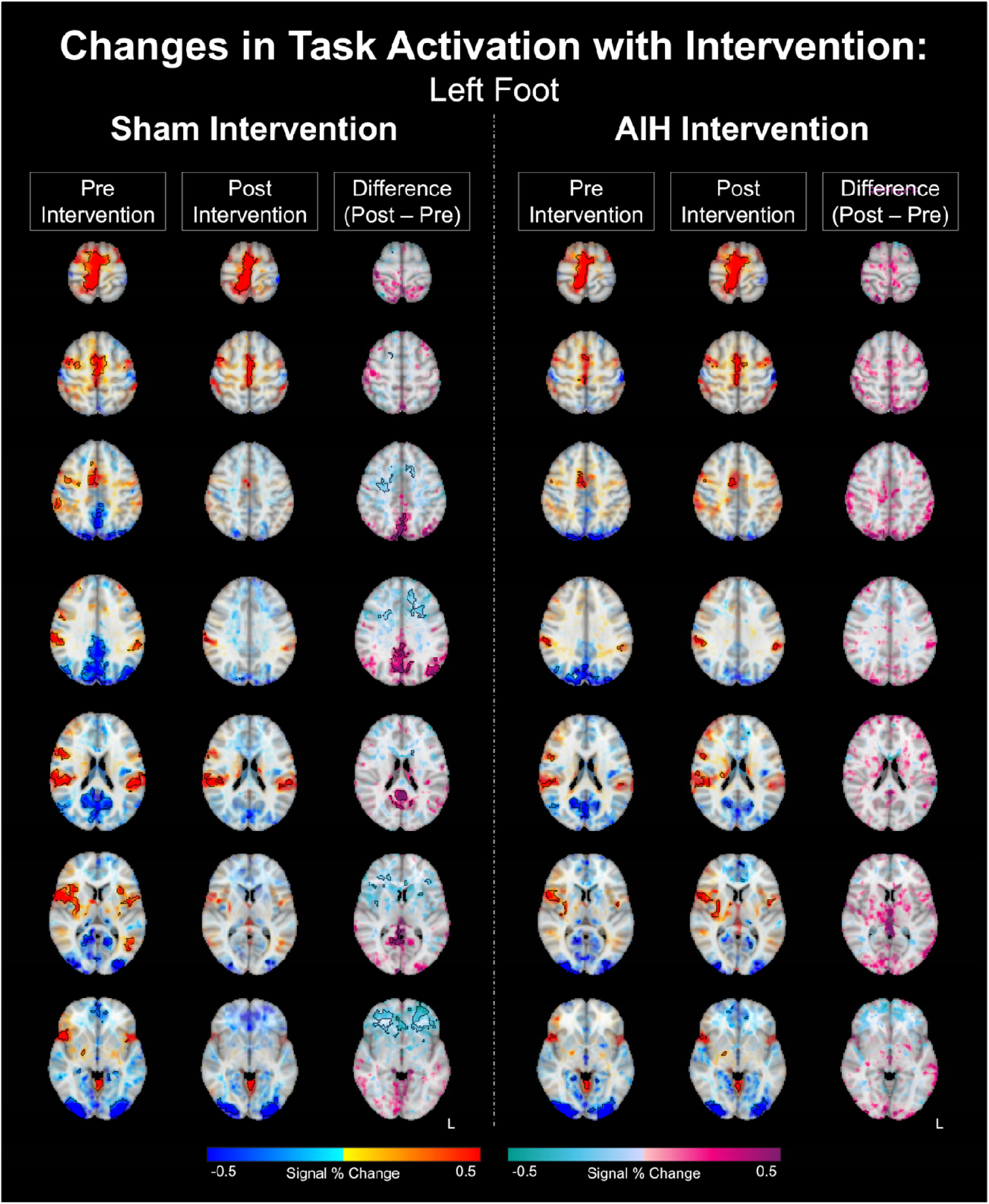
Task Activation at Baseline, Post-Intervention, and Change with Sham and AIH Therapies for the Left Foot. Group maps of pre-intervention and post-intervention activation with the left-foot task (shown with a red-blue colormap) and of changes in activation with intervention (shown with a magenta- teal colormap) were calculated from complete subject datasets (n=21). The opacity of signal changes is quadratically modulated by the associated t-statistic, and clusters of significant activation are outlined in black. A threshold of p<0.001 was used for activation maps and a threshold of p<0.05 was used for difference maps.

Activation maps before and after sham and AIH interventions and the change in activation with each intervention for the right foot are shown in Figure 4. Following the sham intervention arm, activation significantly increased in the precuneate cortex, caused by a decrease in negative BOLD signals. The AIH intervention did not generate any significant increases in activation, although bilateral sub-threshold increases were seen in the opercular cortex similar to the left foot. Increases in negative BOLD signal were seen in the bilateral prefrontal and posterior parietal cortices and the contralateral primary motor and somatosensory cortices, although there was again substantial overlap with white matter (76-95%).

**Figure 4.**
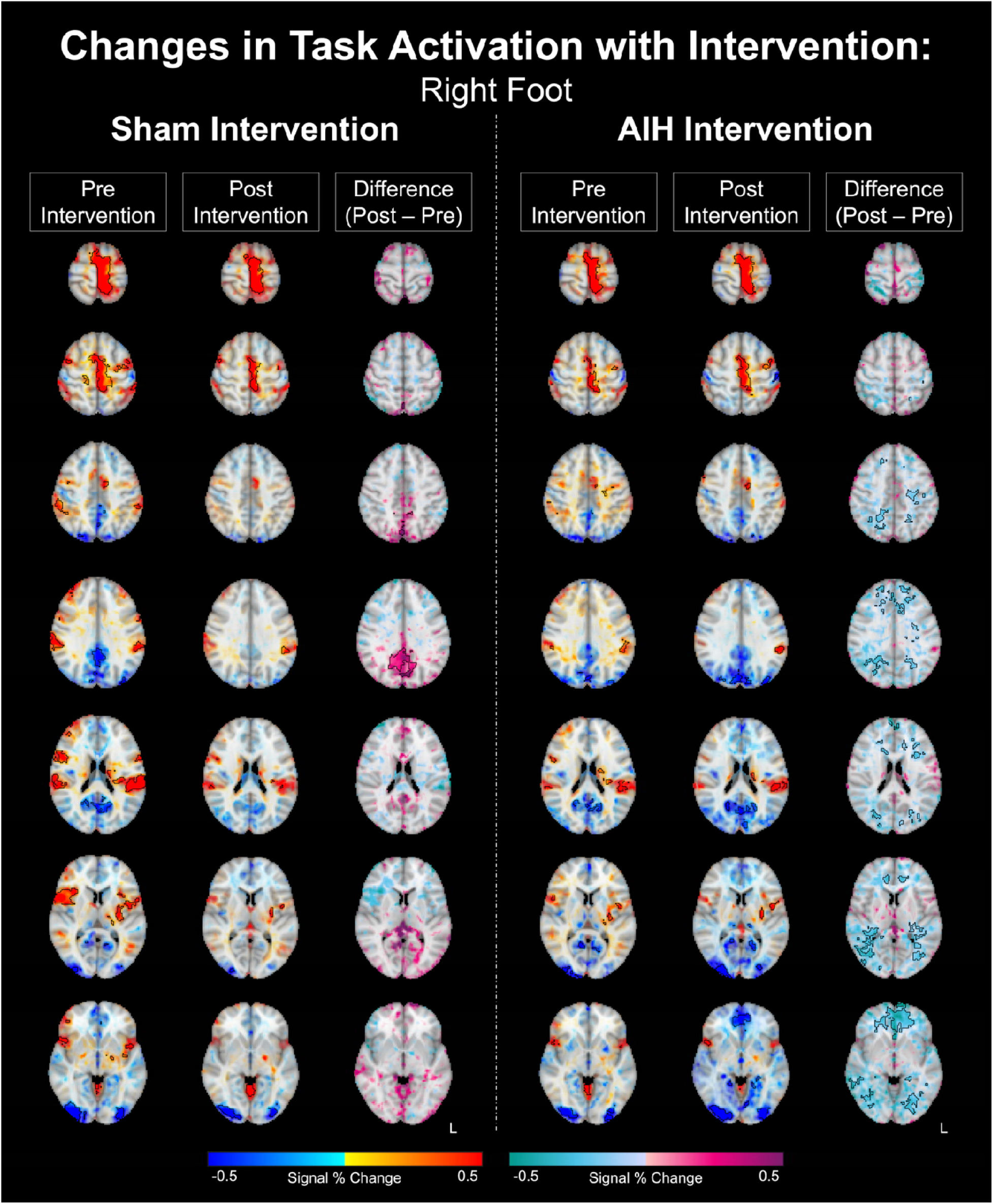
Task Activation at Baseline, Post-Intervention, and Change with Sham and AIH Therapies for the Right Foot. Group maps of pre-intervention and post-intervention activation with the right-foot task (shown with a red-blue colormap) and of changes in activation with intervention (shown with a magenta- teal colormap) were calculated from complete subject datasets (n=21). The opacity of signal changes is quadratically modulated by the associated t-statistic, and clusters of significant activation are outlined in black. A threshold of p<0.001 was used for activation maps and a threshold of p<0.05 was used for difference maps.

Significant changes in task-evoked neural activity with AIH therapy compared to the sham intervention are shown in Figure 5; whole-brain maps are shown in Panel A, whereas detail views of the basal ganglia region, where the primary effect was seen, are highlighted in Panel B and shown against the structures of the caudate, pallidum, putamen, and thalamus. The transparency of activation is modulated by the interaction F-statistic to allow for better consideration of sub-threshold effects. For the left foot, significant increases in BOLD activation were seen bilaterally in the supplemental motor area, the opercular cortex, the insula, and the pallidum and putamen. Significant increases were also seen in the ipsilateral caudate and contralateral thalamus, although sub-threshold increases were also seen in the contralateral and ipsilateral sides, respectively, of these regions and in the contralateral primary motor cortex. No significant decreases were identified for the left foot. For the right foot, no significant increases in BOLD signal activation were seen, although changes approaching significance were again seen bilaterally in the caudate, pallidum, and putamen as well as the contralateral opercular and insular cortices. Significant decreases were seen with right-foot movement in the bilateral prefrontal and posterior parietal cortices and in the ipsilateral superior and middle temporal gyri, all resulting from enhanced negative activation.

**Figure 5.**
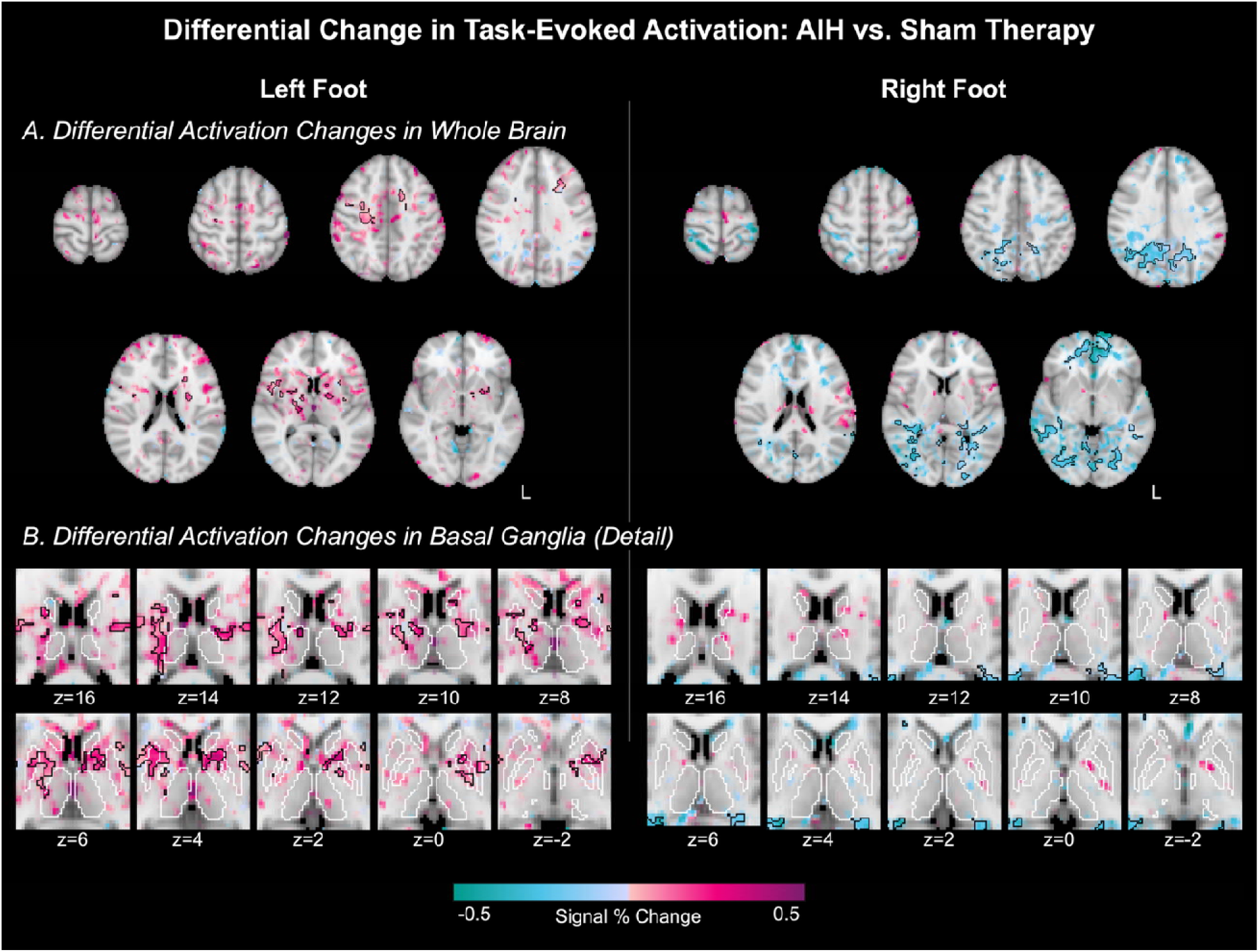
Effect of AIH Intervention on Left- and Right-Foot Task Activation Compared to Sham. Group maps of task activation changes with AIH therapy are shown. The opacity of the signal change is modulated by the interaction (time and intervention) f-statistic and significant interaction clusters, showing where the activation change was significantly different between timepoints and intervention conditions, are outlined in black (p<0.05). Whole-brain maps are shown in Panel A; detail maps of the basal ganglia region with continuous slices are shown in Panel B. Regions of interest (ROIs) of the caudate, pallidum, putamen, and thalamus are outlined in white. ROIs were generated by thresholding the respective regions from the Harvard-Oxford subcortical structural atlas at 75% probability.

## 4 Discussion

This study examined the effects of AIH therapy on neural activation associated with lower limb motor control in PwMS. Contrary to our initial hypothesis, significant decreases in gray matter activation in motor regions were not observed following AIH therapy under this experimental design, suggesting that this AIH therapy did not measurably reverse the compensatory cortical activation patterns that have been identified in PwMS during motor task performance. Instead, the primary effect of AIH therapy in this trial was an increase in activation in the thalamus and basal ganglia regions (putamen, caudate, and pallidum). These changes may reflect a distinct mechanism by which AIH therapy influences lower limb motor function in MS.

### 4.1 Task Activation in Multiple Sclerosis

Motor task fMRI in PwMS probes how the brain maintains motor function despite demyelination and axonal loss. In this study, we examined activation associated with lower-limb movement in PwMS. Lower-limb tasks are less common than upper-limb and hand tasks due to enhanced task-correlated motion artifacts and difficulty isolating movement to the joint of interest, and few previous studies have examined the neural activation patterns associated with lower-limb motor tasks in PwMS^44–47^. Evidence from upper-limb and hand studies suggest that compensatory activation enables patients to maintain relatively normal motor function, and the few studies that have been performed indicate that compensatory activation patterns are also present for lower-limb movement^24,38–40,44–47^.

While no control group was included in the present study, comparisons of the BOLD activation evoked by the lower-limb task in this population with previous research and with expected activation patterns suggests that compensatory activation may be present. Activation was seen in the typical regions activated by this task in healthy controls, i.e. the medial primary and supplemental motor cortices and the primary somatosensory cortex^38–41^. Additional activation was identified bilaterally in the insular, frontal opercular, and parietal opercular (secondary somatosensory) cortices, regions that are commonly implicated in compensatory activation patterns in PwMS^38–41^. The recruitment of additional cortical regions to aid in adequate task performance is unsurprising, given the level of disability in this study cohort and participants’ reports of difficulty performing the task.

### 4.2 Changes in Neural Activation with AIH Therapy

Neural activation showed both significant increases and significant decreases in the AIH intervention arm versus the sham AIH arm (Figure 5). Increases were largely confined to the basal ganglia, insular and opercular cortices, and the primary and supplementary motor and somatosensory regions. BOLD increases were seen bilaterally, with slightly more activation on the side contralateral to the task. Although increases were seen for both feet, significant clusters of increased activation were only found for the left-foot task. Given that the majority of the study cohort showed greater motor and sensory deficits on the left side (Table 1, 12 participants with left-sided weakness, 9 with right-sided weakness), there may be greater potential for improvement on this side. Increases in activation after AIH therapy compared to sham were detected primarily in the deep grey matter structures of the thalamus, putamen, caudate, and pallidum (Figure 5B). These regions comprise key structures of the basal ganglia motor circuit, which coordinates motor control and is particularly involved in the learning, execution, and automation of complex motor functions such as this rhythmic flexion task^48–50^. AIH therapy induces neuroplastic changes by activating serotonergic raphe nuclei, triggering serotonin release and activating serotonin receptors in the neurons to which they project^1,51,52^. Many of these serotonergic projections target the basal ganglia, particularly the caudate and putamen^53^. Increased activation in these regions by the raphe nuclei may explain the observed increases in BOLD signal following AIH therapy, and these enhancements in activation may reflect additional supraspinal mechanisms through which AIH therapy ultimately achieves improvements in motor function.

Decreases were seen in the posterior parietal lobe, the posterior temporal lobe, and the frontal lobe. Frontal lobe hyperactivity has been previously associated with task performance in PwMS, and decreases in activation have been found with previous therapeutic techniques, suggesting increased task network efficiency^54^. However, the posterior parietal and occipital lobe activation changes were primarily confined to white matter, with all clusters exceeding 50% white matter overlap. While white-matter fMRI is increasingly of interest, BOLD signals in white matter are less understood than those in gray matter^55^.

Lower blood flow and weaker hemodynamic responses decrease sensitivity, and the relationship between neural activity and BOLD changes in white matter is not well characterized^55,56^. Furthermore, BOLD signals in white matter will be influenced by the presence of MS lesions, which alter local energy consumption and uncouple metabolic demand from vascular supply^55,57^. Negative BOLD signals in white matter have also been associated with changes to blood and cerebrospinal fluid (CSF) compartment volume, rather than neurovascular coupling mechanisms, particularly in voxels with deep medullary veins or located proximal to the boundary between tissue and CSF, where MS lesions are also frequently located^58^. Thus, while they should not be discounted as purely noise, the observed white matter fMRI changes should therefore be interpreted with caution, particularly at the lower field strength and spatial resolution used in this study. Future studies specifically investigating white-matter fMRI changes in MS could clarify interpretation.

Cerebrovascular changes may also contribute to changes in activation seen with AIH therapy. MS has been associated with cerebrovascular abnormalities, including hypoperfusion and reduced cerebrovascular reactivity, and MS lesions typically form around a central vein, demonstrating that there is a significant vascular component to the disease^59–61^. While changes to whole-brain cerebrovascular function have not previously been seen with AIH therapy in healthy adults, supporting the safety profile of this intervention, we have shown that 12 sessions of AIH enhance focal perfusion to bilateral sensorimotor cortices^62^. The specific cerebrovascular impact of AIH in MS, as well as an understanding of how these changes may influence fMRI activation mapping, will be important targets in future research.

### 4.3 Association of Activation Changes with Fatigue

While fatigue was not measured under the present experimental design, it is likely that our experimental protocol modulated fatigue. The post-intervention scans were performed at the end of a long study visit, after both the AIH therapy and extended functional motor outcome assessments. Anecdotally, participants reported a) feeling tired at the end of the day and b) feeling less tired following what they believed was the true AIH intervention arm (note, the sham AIH is widely accepted to be indistinguishable from the true AIH intervention in practice). Thus, the unintentional fatiguing effects of our experimental design may drive some of the brain changes observed in the sham intervention arm (e.g., reduced deactivation of the precuneus for both left and right foot motor tasks). Furthermore, a major effect of the true AIH intervention may be to simply mitigate those effects, as the precuneus was not significantly different between pre- and post-intervention scans in the AIH arm. The precuneus is part of the DMN, which has previously been implicated in fatigue in PwMS, with individuals experiencing greater fatigue displaying lower functional connectivity of the DMN and basal ganglia^63–65^. However, the observed changes in task- evoked negative signal changes in this study cannot be directly compared to such literature reports of resting state functional connectivity.

Neural activation in the basal ganglia has also been inversely related to the severity of fatigue in neurological disorders^66,67^. Reductions in basal ganglia connectivity and lower activation of the basal ganglia during cognitive tasks have been associated with greater subjective and objective measures of fatigue in multiple sclerosis, suggesting that dysfunction in these regions may be foundational to central fatigue in MS^68,69^. We observed that AIH increased activation of the thalamus, putamen, pallidum, and caudate significantly more than the sham intervention, again suggesting that this intervention may have a beneficial effect on mitigating fatigue. However, measures of fatigue were not obtained in this study, Future work should obtain measures of cognitive and physical fatigue to explore whether this intervention is a viable therapy to modulate central fatigue in MS and other neurological disorders.

### 4.4 Limitations

Limitations of this experimental design present further opportunities for future studies of the effect of therapeutic AIH on motor control in PwMS. This preliminary crossover trial did not include measurements of task performance during the MRI; it is possible that participants’ performance of the ankle flexion task improved with AIH therapy and with practice, and changes in task activation may reflect this improvement. Anecdotal reports from participants suggested that they felt they could perform the task better, and future studies should include quantitative measures of this task performance for incorporation in fMRI analysis to account for this.

This study did not include a concurrent training component. Therapeutic AIH has been found to have a synergistic effect with motor training, priming the motor system to enhance the outcomes that can be obtained from solely AIH therapy or motor training^17^. Although we did observe increased ankle flexion strength in this study following AIH therapy alone (functional outcomes will be reported separately), future trials combining AIH with training will be necessary to evaluate whether the neural changes we have observed relate to meaningful functional improvement (e.g., walking and balance).

The fMRI acquisition was limited in spatial coverage and in spatial resolution. The acquisition did not include the cerebellum in most participants, and it was therefore excluded from analyses. The cerebellum is highly involved in motor control, with particular importance in coordination, balance, and motor learning^70^. Cerebellar pathology is common in MS, causing gait and coordination problems, tremor, and ataxia as well as cognitive problems due to lesions, atrophy, and disruptions to cerebellar connectivity^71–73^. The cerebellum is directly linked to the raphe nuclei, which act as the central command center modulating neuroplasticity, but no study to date has assessed cerebellar changes with AIH therapy^2,74^. Future work should investigate the effects of AIH therapy in the cerebellum. The voxel spatial resolution implemented in this study also did not allow for detailed investigation of the small subcortical regions that showed the greatest changes with the intervention. Future studies of the supraspinal effects of AIH therapy with MRI should implement a smaller voxel size to allow for more accurate localization and co- registration of these salient structures, increasing confidence in analyses of subcortical activation changes.

Finally, MS lesion load, lesion location, and the subsequent motor impairments are highly variable across individuals. Precision mapping in fMRI is becoming increasingly practical with technological and analytical advancements, and single-subject activation in subcortical structures can be robustly detected with approximately 30 minutes of multi-echo fMRI data^75^. Precision mapping approaches would eliminate need to average across the inter-subject variability introduced by MS pathology and could further help to clarify known variability in the efficacy of AIH treatment, as previous studies have reported substantial inter-individual variability in treatment response magnitude^2,76^. Future work should consider applying precision mapping fMRI techniques to studies of supraspinal changes in neural control of movement with AIH therapy.

## 5 Conclusion

Further study of the neuroplastic changes associated with AIH therapy in MS is needed to clarify how the alterations observed here relate to behavioral and functional outcomes. This preliminary crossover trial suggests that neuroimaging may play a valuable role both in identifying candidates likely to respond to AIH therapy and in optimizing AIH protocol parameters to enhance efficacy, positioning basal ganglia activation as a potential candidate biomarker for future trials combining AIH with structured motor rehabilitation.

## 7 Author Contributions

HRJ: Investigation, Formal analysis, Visualization, Software, Validation, Data curation, Writing - Original Draft, Writing - Review & Editing

CZ: Investigation, Formal analysis, Software, Validation, Writing - Review & Editing RT: Investigation, Formal analysis, Software, Validation, Writing - Review & Editing JNN: Investigation, Writing - Review & Editing

NAR: Investigation, Methodology, Software, Writing - Review & Editing AN: Investigation, Writing - Review & Editing

AB: Investigation, Writing - Review & Editing

RK: Project administration, Writing - Review & Editing LO: Investigation, Writing - Review & Editing

BAC: Investigation, Resources, Funding acquisition, Writing – Review & Editing AJN: Investigation, Funding acquisition, Writing – Review & Editing

MS: Conceptualization, Methodology, Resources, Supervision, Funding acquisition, Project Administration, Writing - Review & Editing

MGB: Conceptualization, Methodology, Resources, Supervision, Funding acquisition, Project Administration, Writing - Review & Editing

## 9 Acknowledgements

The authors would like to thank Rachel Young and the Center for Translational Imaging at Northwestern University for supporting data collection.

## Data Availability

All data produced in the present work are available upon reasonable request to the authors.

